# COVID-19 in the Republic of Belarus: pandemic features and the interim safety and efficacy assessment of the Gam-COVID-Vac vaccine

**DOI:** 10.1101/2021.11.15.21265526

**Authors:** Ala M Dashkevich, Veronika S Vysotskaya, Iryna N Hlinskaya, Anzhela L Skuranovich, Aliaksandr A Tarasenka, Inna A Karaban, Natalya D Kolomiets

## Abstract

**Objective:** To study the COVID-19 pandemic features among the population of the Republic of Belarus from February 2020 to September 2021 and assess the safety (tolerance) and epidemiological efficacy of the Gam-COVID-Vac vaccine (Sputnik V).

**Materials and methods:** A retrospective analysis of COVID-19 cases in the Republic of Belarus from the beginning of registration (February 28, 2020) to September 12, 2021 was performed. To assess the COVID-19 case detection dynamics, official registration data available on the website of the Ministry of Health of the Republic of Belarus were used.

Vaccine safety (tolerance) and efficacy were assessed in an observational study. Safety (tolerance) was assessed by presence/absence of adverse reactions: general and local ones.

The efficacy rate (E) and the epidemiological efficacy index (K) was calculated according to the formula: E(%)=100*(b-a)/b, K=b/a.

**Results:** Our data show that The COVID-19 pandemic in the Republic of Belarus is characterized by successive development stages: the first is the absence of COVID-19 cases in the country; the second is the registration of individual infection cases that came from abroad followed by local pathogen spread among the country’s population; the third is a local spread of COVID-19 among individuals who had contact with infected people; the fourth is the detection of cases where patients had no history of exposure to COVID-19 patients.

As of calendar week 26, 2021 Delta variant of SARS-CoV-2 has become the prevalent in the country.

Follow-up results in January-August 2021 showed that the Sputnik V vaccine was well tolerated, with 80,832 adverse reactions reported (2.99% (95% CI 2.9-3.0) of the total number of vaccine doses administered). In terms of severity, adverse reactions were mild (91.4% (95% CI 91.2-91.6)) and moderate (8.6% (95% CI 8.6-8.8)). The epidemiological efficacy rate was 96.3%, the epidemiological efficacy index was 26.7. Thus, the results obtained testify to high prophylactic efficacy of the Sputnik V vaccine.

**Conclusions:** The COVID-19 pandemic in the Republic of Belarus is characterized by successive development stages: from no cases in early 2020 to detected cases where most individuals had no history of contact with COVID-19 patients; periods of rising and falling incidence. The Sputnik V vaccine has demonstrated a high safety profile and epidemiological efficacy throughout mass vaccination in the Republic of Belarus.

## Introduction

The COVID-19 pandemic that emerged in 2020 and still goes on is a global public health challenge. As of September 12, 2021, a total of 224,301,119 cases were registered worldwide according to public sources [1].

In the emerging epidemiological situation, vaccination is an important aspect in countering COVID-19.

## Objective

To study the COVID-19 pandemic features among the population of the Republic of Belarus from February 2020 to September 2021 and assess the safety (tolerance) and epidemiological efficacy of the Gam-COVID-Vac vaccine (Sputnik V).

## Materials and methods

A retrospective analysis of COVID-19 cases in the Republic of Belarus from the beginning of registration (February 28, 2020) to September 12, 2021 was performed. To assess the COVID-19 case detection dynamics, official registration data available on the website of the Ministry of Health of the Republic of Belarus were used.

Coefficients characterizing the epidemic were considered.

The infection reproduction index (R, baseline reproductive number) was calculated based on the number of detected COVID-19 cases over the past 7 days. In general, R > 1 indicates the presence (persistence) of the potential of an infectious disease for epidemic spread in a particular population; R < 1 indicates the absence of such potential [2].

Vaccine safety (tolerance) and efficacy were assessed in an observational study. Safety (tolerance) was assessed by presence/absence of adverse reactions: general (fever, malaise, headache, muscle pain, runny nose, nausea, vomiting, sore throat, etc) and local ones (redness, swelling, soreness at the injection place).

The efficacy rate (E) was calculated according to the formula: E(%)=100*(b-a)/b, where

- E is the efficacy rate,
- a is COVID-19 incidence rate (morbidity) among vaccinated individuals,
- b is COVID-19 incidence rate (morbidity) among unvaccinated individuals

The epidemiological efficacy index (K) was calculated according to the formula: K=b/a, where “a” is disease incidence among vaccinated people, “b” is disease incidence among unvaccinated ones [3].

## Statistical analysis

The data were processed using the Excel 2010 statistical software suite. Confidence intervals (95% CI) were calculated using the method of A.Wald, J.Wolfowitz, corrected by A.Agresti, B.A.Coull [4].

## Results

Under the WHO classification [5], the global COVID-19 pandemic is characterized by 4 development scenarios (stages): the first is the absence of COVID-19 cases in the country; the second is the registration of individual infection cases that came from abroad followed by local pathogen spread among the country’s population; the third is a local spread of COVID-19 among individuals who had contact with infected people; the fourth is the detection of cases where patients had no history of exposure to COVID-19 patients.

The above-mentioned pandemic stages can also be traced in the Republic of Belarus. The first COVID-19 case was detected in the country during calendar week 9 (February 28, 2020): a citizen who arrived from Iran.

Over the following 4 weeks (March 2-29, 2020), mainly sporadic cases were registered, mostly among individuals returning from Europe (Italy, Portugal) and those who had contact with individuals arriving from abroad.

In the initial phase of the epidemic, which lasted for 5 calendar weeks (calendar weeks 9-13, 24.02.-29.03.2020), 1 to 49 COVID-19 cases, with an average weekly incidence rate of 0.2 cases per 100,000 population, were registered. Only one month after the first case was reported (March 30, 2020), 58 new cases per day were detected.

During calendar week 14 (30.03.2020-05.04.2020), local coronavirus spread was noted and the first COVID-19 wave began.

The maximum incidence rate was recorded during calendar week 20 (11.05.2020-17.05.2020) and amounted to 71.4 cases per 100,000 population. The incidence rate peaked on May 17, 2020, when 969 cases were reported.

Thereafter, there was a gradual decrease in the number of cases with a minimum incidence level during calendar weeks 32-33 (9.3 and 6.2 cases per 100,000 population, respectively).

Overall, the first rising incidence period continued in the country for 25 weeks (calendar week 9-33, 24.02.2020-16.08.2020). The total number of cases was 69,516, with an average weekly incidence rate of 29.7 cases per 100,000 population (Figure 1).

**Figure. 1.**
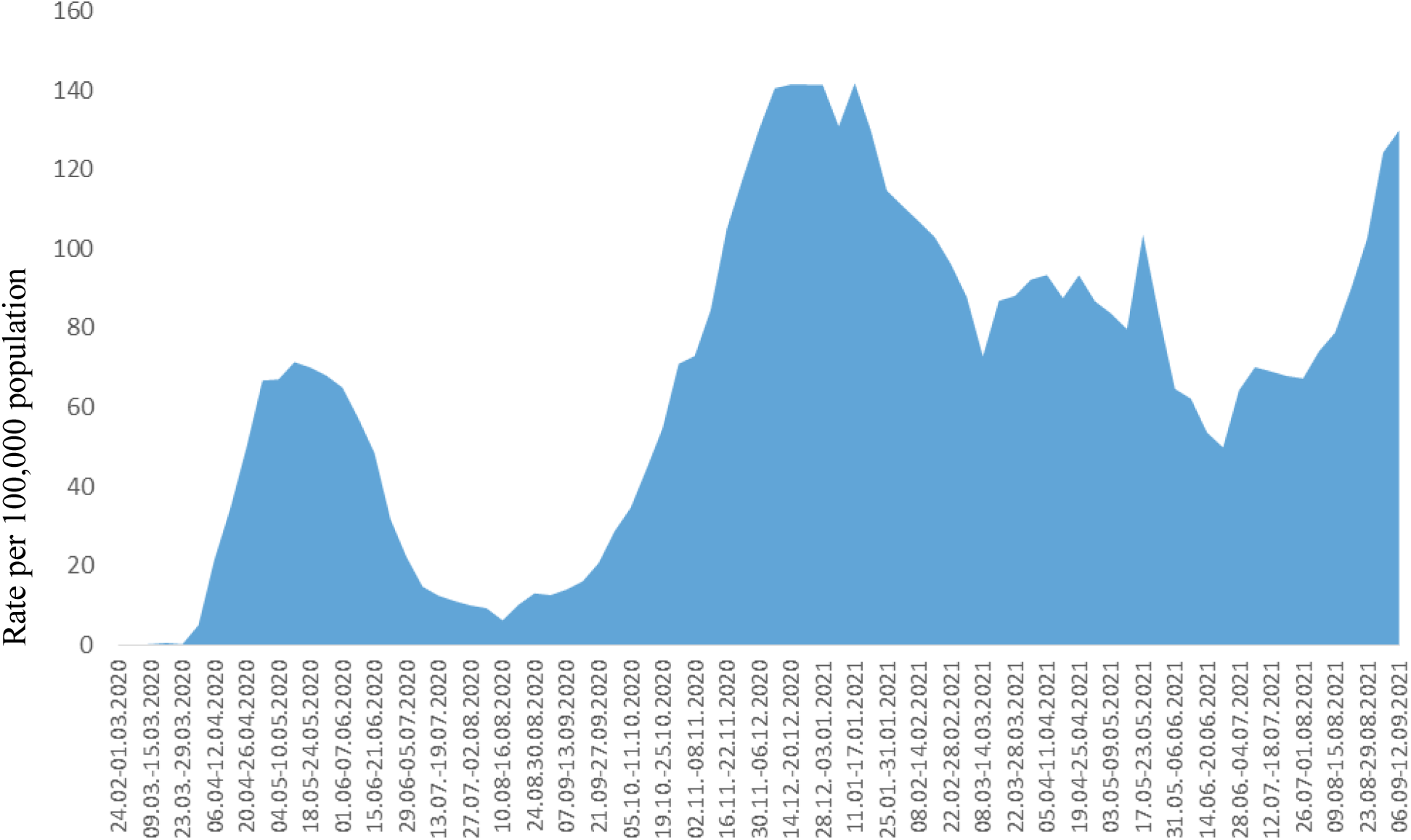
Dynamics of the incidence of COVID-19 in the population of the Republic of Belarus in 2020-2021

As of calendar week 34, an upward trend began with a rapid increase in the number of cases. The maximum incidence rate was recorded on calendar week 2, 2021 (11.01.2021-17.01.2021): 141.8 cases per 100,000 population and was 1.98 times higher than the maximum rate of the first rising incidence period. The peak incidence was recorded on January 13, 2021: 1,972 COVID-19 cases, which is twice higher than the previous peak incidence.

Overall, the second, more intense and prolonged, rising incidence period lasted for 30 calendar weeks: until calendar week 10, 2021 (08.03-14.03.2021). The average weekly incidence rate increased 2.8 times compared to the first rising incidence period and amounted to 83 cases per 100,000 population. During this period, COVID-19 cases were noted for which no epidemiological link could be established between patients (i.e. patients had no history of contact with COVID-19 patients or other respiratory infections).

Molecular genetic analysis of SARS-CoV-2 isolates collected in Belarus from February 2020 to February 2021 showed the spread of 4 COVID-19 clades in the country: O (5%), G (25%), GR (27.5%), GH (40%) and GRY (2.5%). A wide representation of the Pangolin lineage (52.5% of the genomes belonged to lineage B.1.) indicates that the SARS-Cov-2 virus came to Belarus from various areas [6].

During the next 4 calendar weeks (March 15, 2021 to April 11, 2021), another upward trend was observed, with daily cases ranging from 876 to 1,651. During calendar weeks 15-25, COVID-19 incidence showed a decreasing trend.

As of calendar week 26, 2021, another rise in the incidence was recorded, due to the spread of the Delta SARS-CoV-2 variant in Belarus. Delta variant of SARS-CoV-2 has since become the prevalent in the country.

A total of 503,073 COVID-19 cases were reported from February 28, 2020 to September 12, 2021, of which 493,053 ended in recovery. The cumulative incidence rate was 5,380.7 per 100,000 population.

The first COVID-19 fatality was registered on March 31, 2020, 6 weeks after the first case was detected. As of September 12, 2021, 3,917 patients, or 0.8% of the total number of cases, died.

An important indicator of the COVID-19 pandemic development intensity is the infection spread index (R).

At the initial stage of the COVID-19 pandemic, the infection spread index (R) was characterized by marked variability (from 0.3 to 26), which probably determined a rapid infection spread.

During the first rising incidence period, R ranged from 0.7 to 1.6; during the second one, R ranged from 0.8 to 1.3. Since August this year, R has been above 1.

As of December 2020, vaccination has been deployed nationwide: health workers, as the top occupational risk group, were the first to be vaccinated.

The National Plan on Vaccination against COVID-19 for 2021-2022 approved by the government of the Republic of Belarus defines immunization tactics including 4 stages of sequential inclusion of specific groups in the vaccination campaign covering 60% of the population.

The first 3 stages provide for immunization of individuals in various risk groups: healthcare and pharmaceutical workers; social care and educating institution staff; individuals aged 61 and older and those with chronic diseases; other individuals with a higher risk of infection and a more serious course of the disease. Then the rest of the adult population are to be vaccinated.

2 vaccines are used for immunization: the vector vaccine Gam-COVID-Vac or Sputnik V (Russian Federation) and the inactivated vaccine against SARS-CoV-2 (Vero Cell) by Sinopharm/BIBP (PRC). Vaccination was carried out in accordance with the instruction for medical use.

As of September 1, 2021, 1,661,524 Belarusians received at least the first shot, of whom 1,374,677 were fully vaccinated.

Full vaccination against COVID-19 was provided to 79.3% of healthcare workers; 64.3% of workers in 24-hour institutions; 46.2% of workers in educational institutions; 26.7% of individuals aged over 60 and those with chronic diseases.

The largest number of vaccinated individuals received Sputnik V: 1,486,517 received a single shot and 1,215,643 got both.

The Sputnik V vaccine is a combined vector 2-component vaccine consisting of a recombinant human adenovirus type 26 (rAd26) vector and a recombinant human adenovirus type 5 (rAd5) vector, both components carrying the SARS-CoV-2 full-length protein S gene [7, 8]. Each vaccine dose contains (1.0±0.5)×10^11^ per dose of each of the recombinant adenoviruses; 0.5 ml per dose for intramuscular injection [9].

Follow-up results in January-August 2021 showed that the Sputnik V vaccine was well tolerated, with 80,832 adverse reactions reported (2.99% (95% CI 2.9-3.0) of the total number of vaccine doses administered). In terms of severity, adverse reactions were mild (91.4% (95% CI 91.2-91.6)) and moderate (8.6% (95% CI 8.6-8.8)). The most frequent adverse reactions included injection place pain, hyperthermia and asthenia. These results are consistent with those of the randomized controlled phase 3 trial in the Russian Federation [8].

Serious adverse events after administering component I of the Sputnik V vaccine were reported in 2 patients in the age groups 30-35 years and 50-54 years: diagnoses “Allergic urticaria following the drug administration”, ICD-10 code L 50.0; “Anaphylactoid allergic reaction to the administration of Gam-COVID-Vac*”*, ICD-10 code T.88.6. Both patients fully recovered without consequences.

According to the preliminary research, these reactions were caused by individual reactions to an adequately prescribed and correctly administered medicinal product. The conclusion about the relation to the vaccination will be available after the corresponding investigation.

During the period in question, there were 281 deaths among vaccinated population (irrespective of the cause of death) and none of them were related to vaccination.

The incidence rate of COVID-19 cases (morbidity) among vaccinated population was 0.19% and the incidence rate (morbidity) among unvaccinated individuals was 5.08%.

The epidemiological efficacy rate was 96.3%, the epidemiological efficacy index was 26.7. Thus, the results obtained testify to high prophylactic efficacy of the Sputnik V vaccine.

## Conclusions

1. The COVID-19 pandemic in the Republic of Belarus is characterized by successive development stages: from no cases in early 2020 to detected cases where most individuals had no history of contact with COVID-19 patients; periods of rising and falling incidence.
2. The Sputnik V vaccine has demonstrated a high safety profile throughout mass vaccination in the Republic of Belarus. The identified adverse reactions were considered as mild to moderate and did not require medical intervention.
3. Serious adverse events after vaccination of Sputnik V were registered only in 2 individuals. Both recovered without consequences. The conclusion about relation to the vaccination will be available after the corresponding investigation.
4. The vaccine demonstrated its epidemiological efficacy. During the 7-8 months of follow-up, the incidence in the vaccinated was 96.3% lower than in the unvaccinated.

Further data collection would permit specifying efficacy in various age groups and among those vaccinated at different time points from the start of the nationwide immunization campaign.

## Data Availability

All data produced in the present study are available upon reasonable request to the authors

## Notes

### Competing Interest Statement

The authors have declared no competing interest.

### Funding Statement

This study did not receive any funding

### Author Declarations

Ethics committee of State Educational Establishment Belarusian Medical Academy of Postgraduate Education gave ethical approval for this work

